# Multi-Night Home testing with portable EEG shows Improvement in Sleep Quality after Starting CPAP for Obstructive Sleep Apnoea

**DOI:** 10.1101/2025.01.23.25320588

**Authors:** Kieran O. Lee, Tristan A. Bekinschtein, Ian E. Smith

## Abstract

Obstructive sleep apnoea (OSA) impacts sleep quality and has considerable night to night variability. This study explored the use of low-density EEG headbands to evaluate changes in sleep architecture and sleep features in the patients’ own homes over multiple nights both Pre and Post treatment with continuous positive airway pressure (CPAP). Seven participants recorded 144 high-quality EEG sessions (84 Pre-CPAP, 60 Post-CPAP). Comparing Pre and Post-CPAP there were significant reductions in N1 (P < 0.0001) and N2 (P < 0.0001) sleep, increases in N3 (P < 0.0001) and REM (P < 0.0001) sleep and fewer sleep stage shifts (P < 0.0001), micro-arousals (P < 0.0001) and awakenings (P < 0.0001). Total sleep time did not change. Strength testing identified that one participant (who had the most severe OSA, and produced the most EEG recordings), was a major driver of the effects, however, improvements in REM, shifts, and micro arousals remained when this participant was excluded. These results suggest low-density EEG devices can be used for multi-night monitoring, potentially advancing OSA research and personalised care.

**Statement of Significance:** OSA is a highly prevalent condition, with increasing global demand for care resources. Current OSA diagnostic and follow-up methods face two significant limitations: they do not capture night-to-night variability in sleep quality, and polysomnography, the current gold-standard measurement of sleep quality, requires substantial resources and is usually performed in a laboratory, which will not always reflect sleep quality achieved at home. This study collected 144 high-quality sleep recordings from seven participants using low-density EEG headbands in a community setting, both Pre and Post-CPAP. Results showed significant improvements in multiple measures of sleep quality after initiating CPAP. These findings demonstrate the feasibility of portable EEG headbands for OSA and suggest potential for enhancing clinical monitoring and advancing research through multi-night frameworks.

## Introduction

OSA has a detrimental effect on sleep quality which can cause daytime symptoms including excessive sleepiness and attentional deficits. CPAP is effective in restoring many sleep features. The largest published study of sleep EEG and cognitive performance after CPAP treatment for OSA assessed 167 patients ^1^. Six months of CPAP use was associated with a mean reduction in N2 of 60 minutes and 59 and 21 minute increases of N3 and REM (rapid eye movement) sleep respectively. The total sleep time did not change. There were positive associated changes in working memory and set-shifting. These changes did not occur in 28 patients using sham CPAP over a 12 week period. Similar, smaller studies have shown effects across all sleep stages, and provided cross-demographic consistency ^2,3^.

Despite the strong indication that CPAP is effective in restoring undisrupted sleep, these studies have a significant limitation in that they only use a single night’s assessment, which has been shown to be a problem when assessing the quality of sleep in healthy participants ^4,5^ and within OSA care and research ^6–8^ due to a high degree of night-to-night variation.

The high global prevalence ^9^, and diverse range of OSA phenotypes ^10^, mean that much of the research conducted on EEG response to CPAP may not be applicable to subgroups of OSA such as patients with only mild OSA. For example a multi-night study by Parrino et al included only males with severe OSA ^11^. If OSA medicine is to move towards a more personalised care program ^12^, there needs to be a practical and affordable method to conduct multi-night assessments across the OSA diagnostic and early treatment monitoring period.

Polysomnography is the gold-standard measurement of sleep quality but is resource-intensive^13^. Here we have used low-density EEG headbands during a standard OSA care pathways to test if it is possible to identify changes in sleep quality Pre-CPAP and Post-CPAP in the home setting. If this method is robust enough to detect neurological changes, as previously demonstrated in polysomnography studies, there is a broad potential to utilise portable EEG monitoring equipment for both research and clinical purposes.

## Methods

### Study Design and Participants

This study used an observational design to monitor the sleep quality of patients attending Royal Papworth Hospital NHS Foundation Trust, Sleep Centre, Cambridge, UK, as they became diagnosed with OSA and started CPAP therapy. Potential participants who had been referred to the Centre with suspected OSA were invited to take part in a study assessing multi-night sleep quality at home by advertisement sent alongside their initial appointment letter. Eligible and willing participants provided informed consent remotely via the online platform DocuSign. At the time of enrolment participants had not yet undertaken an OSA screening test or had a consultation with a secondary care physician for their current referral. Patients were not recruited if they had used CPAP therapy within the last 6 months. Initially participants were asked to make nightly sleep recordings, for as many nights as possible, pending a diagnostic decision. Diagnoses were made using at-home pulse oximetry, respiratory polygraph or polysomnography, depending on the patient’s circumstance. If OSA was diagnosed and a decision was made for a trial of CPAP therapy, a further 2 weeks of monitoring were requested after treatment had started. This method facilitated collecting Pre-CPAP and Post-CPAP data without having any influence of care provision or time of care delivery. Ethical approval for the study was granted by the UK Health Research Authority on the 30th of May 2023 (IRAS Project ID: 314762).

### Equipment

Consenting patients were provided with a Dreem 2 or 3 portable low-density EEG headband, to use at home when sleeping. These are commercially available devices with a seven electrode configuration (3 frontal, 2 occipital), to produce seven channels of EEG signal. Dreem has an automatic sleep stage and sleep features scoring algorithm, which provides data on sleep stage duration and percentage, shifts (number of sleep stage changes), micro arousals (abrupt shift of EEG frequency, greater than 16 Hz lasting between 3 and 15 seconds), awakenings and respiration. The automated scoring has been validated against polysomnography ^14^ and has been used in other sleep experiments ^15–17^. As a precautionary measure we excluded all recordings which had a REM proportion greater than 45%, due to some evidence suggesting REM sleep can be overestimated by the Dreem headband ^18^, likely because Dreem does not collect electromyography (EMG) signal, which is associated with more accurate REM classification ^19^. All participants were given a training session on how to correctly use the headband, and were provided with the study team’s contact details. They were encouraged to reach out if any issues arose during the monitoring period.

### Data retention and statistical analysis

We collected 539 community based EEG sleep recordings with more than 4 hours total sleep time from 35 participants screened for OSA between August 2023 and August 2024. Seventeen participants were not used for the current analysis due to a negative diagnostic test, or did not initiate a course of CPAP (5 had a confirming test but were treated with a mandibular advancement device as opposed to CPAP). This left 408 recordings from 19 participants who started CPAP. One participant was excluded due to abnormally high scorings of REM and having a prior history of a traumatic brain injury. Additionally, recordings failing Dreem’s strict record quality threshold (30% of recordings), or with REM proportion’s greater than 45% were excluded (at an equal percentage loss for both Pre-CPAP and Post-CPAP recordings), resulting in a total session loss of 35%. The final dataset included 215 EEG recordings from 13 participants for confidence testing, and 144 from 7 patients (84 Pre-CPAP, 60 Post-CPAP) for primary analysis. Due to the opportunistic nature of the design, varying quantities of data were collected across participants and conditions and between individuals. To address this we performed mixed-effects modelling to examine the influence of treatment conditions (Pre-CPAP and Post-CPAP) on sleep features of interest (fixed effects), with individual participants set as random factors to minimise the impact of individual differences and sample variability. All analysis was conducted in MATLAB (version R2024a).

## Results

### Group Results

Seven participants (2 female) recorded retainable data in both Pre-CPAP and Post-CPAP treatment conditions: mean age of 63 (range: 36 – 76) years, BMI: 40.7 (range: 25.5 – 46.1) kg/m^2^, and self-reported weekly alcohol consumption of 2.5 (range 0 – 8) units. None of the participants were on regular prescription sedatives or stimulants, two were taking antidepressants. The mean Oxygen Desaturation Index / Apnoea Hypopnea Index at diagnosis was 35.7 (range 5.9 – 86.7), at a split of 4 severe cases, 1 moderate and 2 mild. The mean Epworth Sleepiness Scale score was 15.6 (range 8 – 21). During the first 2 weeks of CPAP there was a group average compliance of 8 hours 2 mins per night at a pressure of 12.4 (range 6.6 – 15.9) hPa. Five participants averaged above 6 hours usage per night, both for the entire 2 week duration and nights EEG was recorded. One patient averaged less than 6 hours and one patient averaged less than 3 hours, although this rose to more than 5 hours on nights when a successful EEG recording was made. Table 1 provides a detailed breakdown of individual participant demographics and characteristics.

**Table 1:**
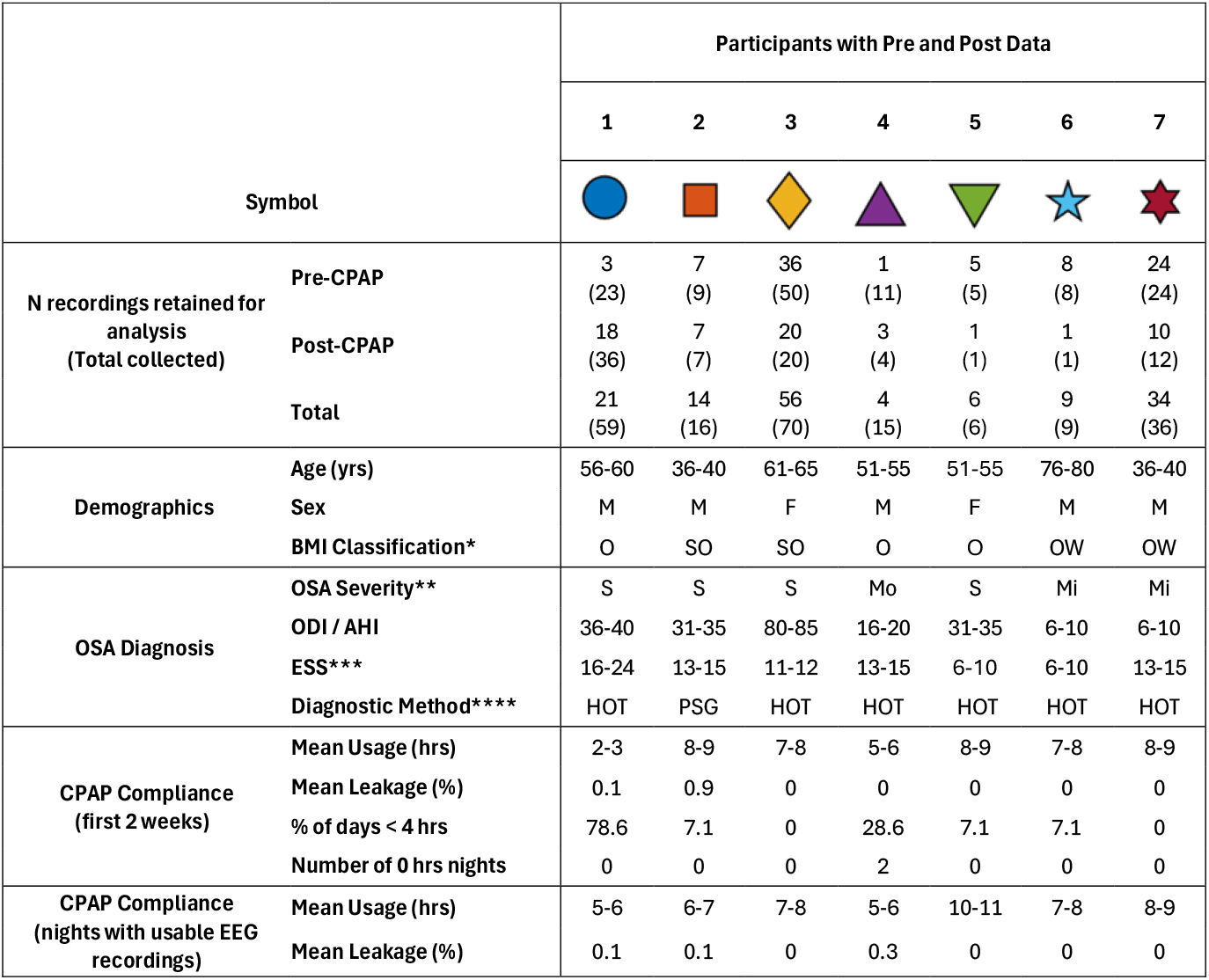
Participant Demographics and Characteristics. The demographic and clinical characteristics of each study participant who contributed usable data in both treatment conditions. Ranges have been applied to protect confidentiality. *OW = Overweight, O = Obese, SO = Severely Obese, **Mi = Mild, Mo = Moderate, S = Severe, ***0-5 = Lower Normal Daytime Sleepiness, 6-10 = Higher Normal Daytime Sleepiness, 11-12 = Mild Excessive Daytime Sleepiness, 13-15 = Moderate Excessive daytime Sleepiness, 16-24 = Severe Daytime Sleepiness, ****HOT = Home Oximetry Testing, PSG = full in lab Polysomnography.

| Symbol |  | Participants with Pre and Post Data |  |  |  |  |  |  |
| --- | --- | --- | --- | --- | --- | --- | --- | --- |
|  |  | 1 | 2 | 3 | 4 | 5 | 6 | 7 |
| **N recordings retained for analysis (Total collected)** | **Pre-CPAP** | 3<br>(23) | 7<br>(9) | 36<br>(50) | 1<br>(11) | 5<br>(5) | 8<br>(8) | 24<br>(24) |
|  | **Post-CPAP** | 18<br>(36) | 7<br>(7) | 20<br>(20) | 3<br>(4) | 1<br>(1) | 1<br>(1) | 10<br>(12) |
|  | **Total** | 21<br>(59) | 14<br>(16) | 56<br>(70) | 4<br>(15) | 6<br>(6) | 9<br>(9) | 34<br>(36) |
| **Demographics** | **Age (yrs)** | 56-60 | 36-40 | 61-65 | 51-55 | 51-55 | 76-80 | 36-40 |
|  | **Sex** | M | M | F | M | F | M | M |
|  | **BMI Classification\*** | O | SO | SO | O | O | OW | OW |
| **OSA Diagnosis** | **OSA Severity\*\*** | S | S | S | Mo | S | Mi | Mi |
|  | **ODI / AHI** | 36-40 | 31-35 | 80-85 | 16-20 | 31-35 | 6-10 | 6-10 |
|  | **ESS\*\*\*** | 16-24 | 13-15 | 11-12 | 13-15 | 6-10 | 6-10 | 13-15 |
|  | **Diagnostic Method\*\*\*\*** | HOT | PSG | HOT | HOT | HOT | HOT | HOT |
| **CPAP Compliance (first 2 weeks)** | **Mean Usage (hrs)** | 2-3 | 8-9 | 7-8 | 5-6 | 8-9 | 7-8 | 8-9 |
|  | **Mean Leakage (%)** | 0.1 | 0.9 | 0 | 0 | 0 | 0 | 0 |
|  | **% of days < 4 hrs** | 78.6 | 7.1 | 0 | 28.6 | 7.1 | 7.1 | 0 |
|  | **Number of 0 hrs nights** | 0 | 0 | 0 | 2 | 0 | 0 | 0 |
| **CPAP Compliance (nights with usable EEG recordings)** | **Mean Usage (hrs)** | 5-6 | 6-7 | 7-8 | 5-6 | 10-11 | 7-8 | 8-9 |
|  | **Mean Leakage (%)** | 0.1 | 0.1 | 0 | 0.3 | 0 | 0 | 0 |

Group analysis showed that comparing Pre and Post-CPAP, total sleep time did not change (P = 0.58, Cohen’s d = 0.11). However there were substantial changes in sleep stage distributions, with the mixed effect model returning reductions respectively of 26.7 and 55.4 minutes for N1 (P < 0.0001, Cohen’s d = -1.44), and N2 (P < 0.0001, Cohen’s d = -1.11), and increases of 35.6 and 53.6 minutes in N3 (P < 0.0001, Cohen’s d = 1.17), and REM (P < 0.0001, Cohen’s d = 1.62) with CPAP. Decreases were found for micro arousals by 27.4 per hour (P < 0.0001, Cohen’s d = - 2.11), awakenings by 18 per night (P < 0.0001, Cohen’s d = -1.36) and sleep stage shifts by 73.6 per night (P < 0.0001, Cohen’s d = -1.64). There were increases in mean respiratory rate over the whole recording by 0.4 bpm (P < 0.001, Cohen’s d = 0.65), 0.2 during N1 (P < 0.08, Cohen’s d = 0.34), 0.4 during N2 (P < 0.001, Cohen’s d =0.74), and 1 in REM (P < 0.0001, Cohen’s d = -1.39). Respiration during N3 showed a marginally significant decrease of 0.3 bpm (P =0.04, Cohen’s d = - 0.4), and awake respiration was not different across conditions. See table 2 for the full list of statistical results. For reliability, all results were re-tested using the entire cohort of usable data (215 EEGs from 13 participants), which included participants who were only able to record usable data during the Pre-CPAP condition. All results and significance were the same, except for respiratory rate in N2 and N3 which were no longer different. Figure 1, shows violin plots of overall distribution of data in both conditions, as well as mean change for each individual.

**Table 2:** Statistical Results.

|  |  | Estimate | SE | tStat | DF | pValue | Cohen's d |
| --- | --- | --- | --- | --- | --- | --- | --- |
| **Sleep Stage Duration (Minutes)** | **N1 Duration** | -26.7 | 3.4 | -7.8 | 142 | < 0.0001 | -1.44 |
|  | **N2 Duration** | -55.4 | 9.3 | -6.0 | 142 | < 0.0001 | -1.11 |
|  | **N3 Duration** | 35.6 | 5.6 | 6.3 | 142 | < 0.0001 | 1.17 |
|  | **REM Duration** | 53.6 | 6.1 | 8.8 | 142 | < 0.0001 | 1.62 |
| **Sleep Stage Proportion (% of TST)** | **N1 Percentage** | -7.1 | 0.9 | -7.9 | 142 | < 0.0001 | -1.48 |
|  | **N2 Percentage** | -14.8 | 1.7 | -8.5 | 142 | < 0.0001 | -1.57 |
|  | **N3 Percentage** | 8.7 | 1.4 | 6.1 | 142 | < 0.0001 | 1.13 |
|  | **REM Percentage** | 13.1 | 1.3 | 9.8 | 142 | < 0.0001 | 1.80 |
| **Sleep Features** | **Total Sleep Time (Minutes)** | 6.1 | 10.5 | 0.6 | 142 | 0.56 | 0.11 |
|  | **Micro Arousals (Per Hour)** | -27.4 | 2.4 | -11.3 | 142 | < 0.0001 | -2.11 |
|  | **Awakenings (Per Night)** | -18.0 | 2.5 | -7.3 | 142 | < 0.0001 | -1.36 |
|  | **Sleep Stage Shifts (Per Night)** | -73.6 | 8.3 | -8.9 | 142 | < 0.0001 | -1.64 |
| **Respiration (BPM)** | **Average total** | 0.4 | 0.1 | 3.5 | 142 | < 0.001 | 0.65 |
|  | **Awake average** | 0.2 | 0.1 | 1.8 | 142 | 0.08 | 0.34 |
|  | **N1 average** | 0.4 | 0.1 | 3.9 | 142 | < 0.001 | 0.74 |
|  | **N2 average** | 0.2 | 0.1 | 2.3 | 142 | 0.02 | 0.43 |
|  | **N3 average** | -0.3 | 0.1 | -2.1 | 131 | 0.04 | -0.40 |
|  | **REM average** | 1.0 | 0.1 | 7.4 | 141 | < 0.0001 | 1.39 |

**Figure 1:**
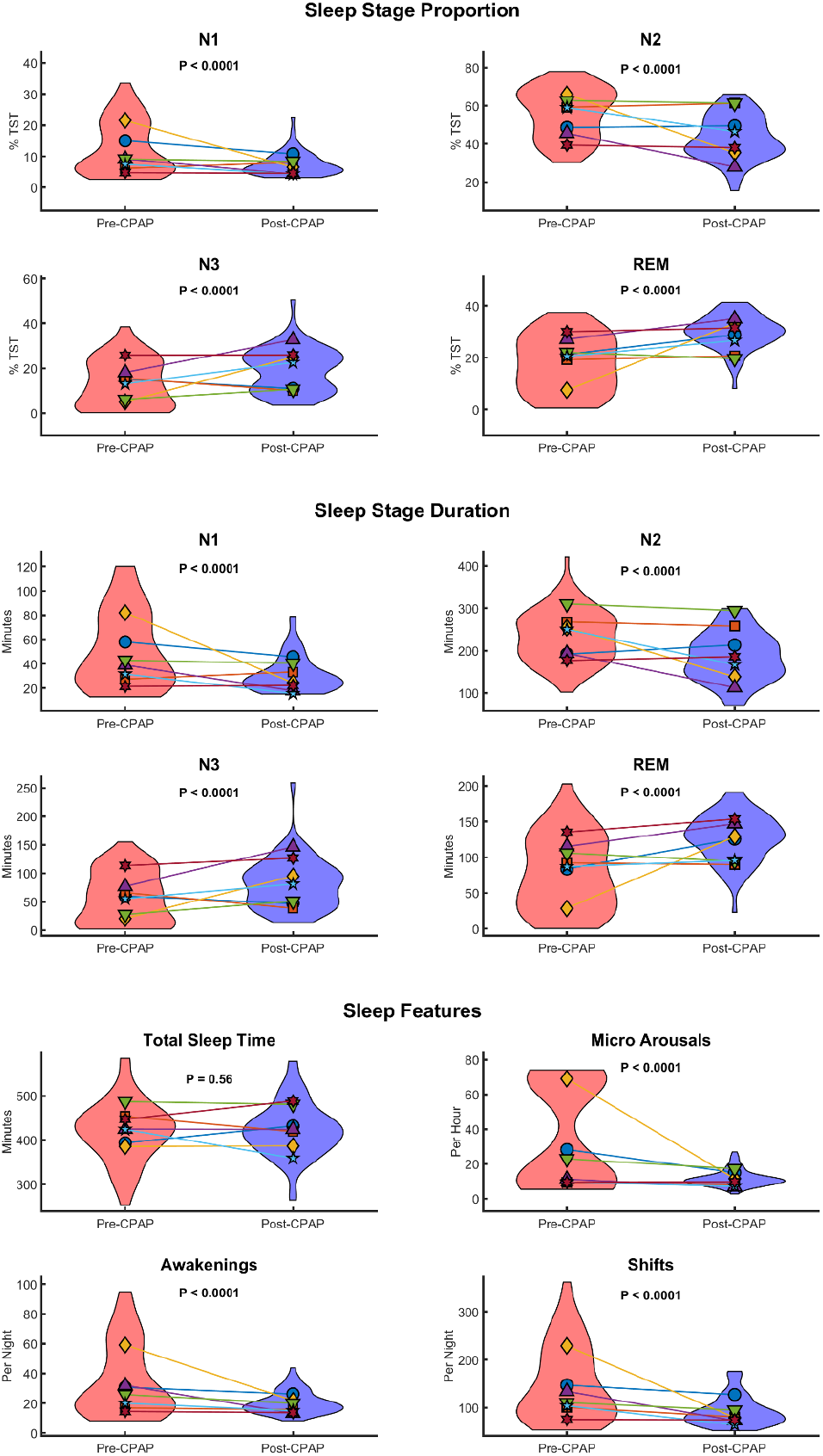
Violin plots showing the distribution of all EEG sessions Pre-CPAP (red) and Post-CPAP (blue). Coloured shapes / lines reflect the mean values of individual participants for each sleep measurement. See Table 1 for colour / symbol key with demographics and characteristics of each participant.

### Individual Results and strength testing

Due to the design and methodology of our study, we retained an unbalanced number of observations across conditions, with the most active participant producing 56 EEG recordings (36 Pre-CPAP / 20 Post-CPAP), and the least active producing 4 (1 Pre-CPAP / 3 Post-CPAP). A degree of variation in data samples was expected and we used mixed effect modelling to control for this, however because the patient with the highest OSA diagnostic result (ODI: 84.2), also contributed the most night recordings we performed additional strength testing by re-calculating p and Cohen’s d effect sizes for each measure with each single individual removed for robustness. For sleep stage duration, sleep stage proportion and sleep features there was no difference in direction of effect or significance, for any iteration when participants 1,2,4,5,6 or 7 were removed. When participant 3 was removed REM duration (P =0.03, Cohen’s d = 0.54), REM proportion (P = 0.02, Cohen’s d = 0.58), micro arousals (P < 0.01, Cohen’s d = -0.7), and shifts (P = 0.02, Cohen’s d = -0.62), all retained significance and directionality of the main result, albeit with a smaller effect size. All other sleep stage proportions and durations, as well as awakenings (marginal: P = 0.05, Cohen’s d = -0.48) were no longer significant. But the directionality of the effect size was consistent with the main effect for all non-significant results, except for N3 proportion (P < 0.86, Cohen’s d = -0.04), N3 duration was (P =0.51, Cohen’s d = 0.16). Figure 2 shows a heatmap of the strength of each sleep measure when any one of participants was removed from the sample.

**Figure 2:**
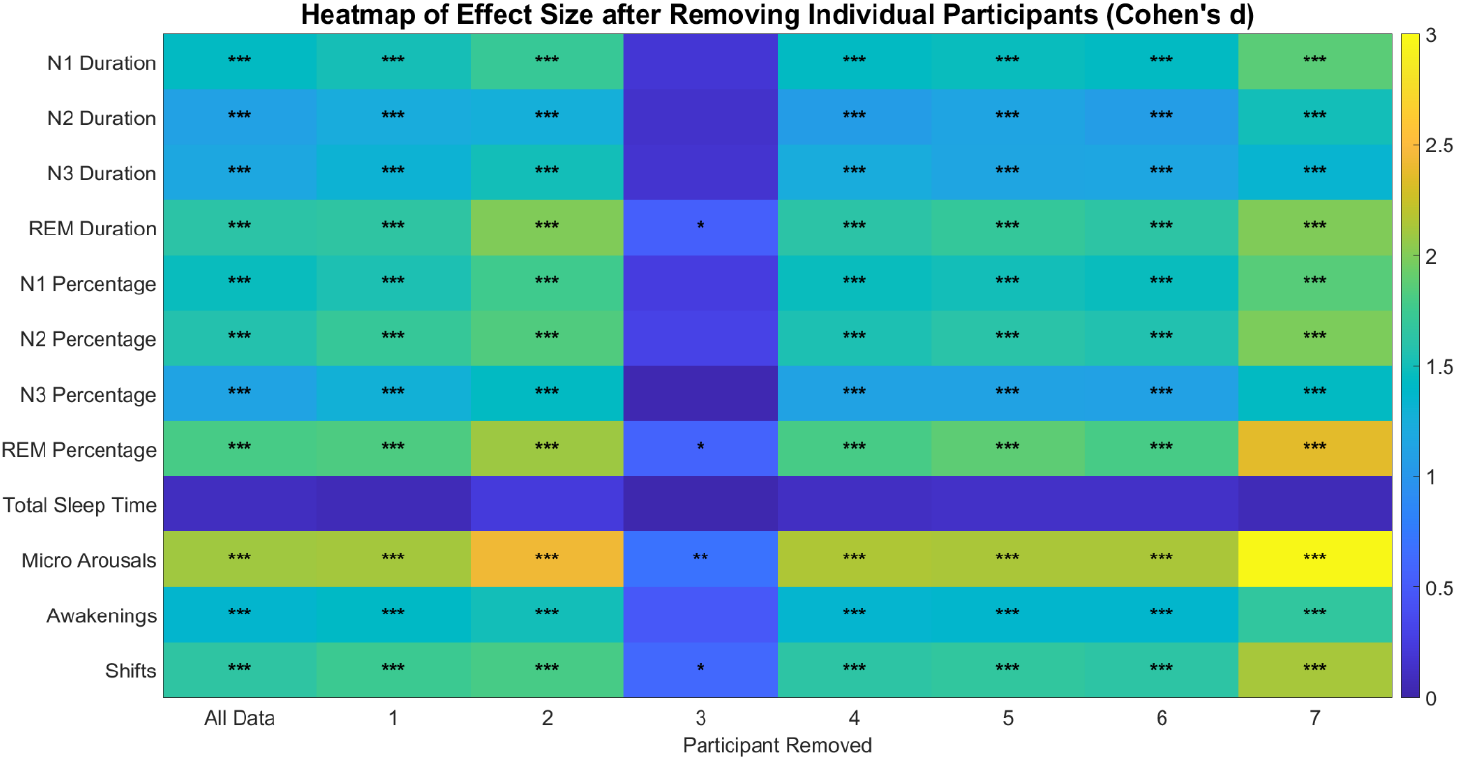
Heatmap of absolute value of effect size for each measure of interest with all, and each individual participant removed, * = P <0.05, ** = P < 0.01, *** = P < 0.0001

### Multi-Night sleep assessment in an individual

The large number of high-quality recordings produced by participant 3, provides a unique opportunity to assess deviations in sleep quality over more than nine weeks during CPAP initiation. Figure 3 shows this participant’s nightly sleep stage durations and proportions in relation to CPAP initiation. There is a clear change in sleep architecture from the first night of CPAP use, with increases in REM and N3 sleep and decreases in N2 and N1 sleep (consistent with our main finding), which remain stable across subsequent nights on treatment. Pre-CPAP data show greater variation across nights. For example, recordings from 49, 36, and 4 days before CPAP show a reasonable amount of N3 sleep. In contrast, recordings from 35, 34, 33, 30, 25, 23, 21, 19, 18, 15, 14, 11, 7, and 2 days before CPAP show little or no N3 sleep.

**Figure 3:**
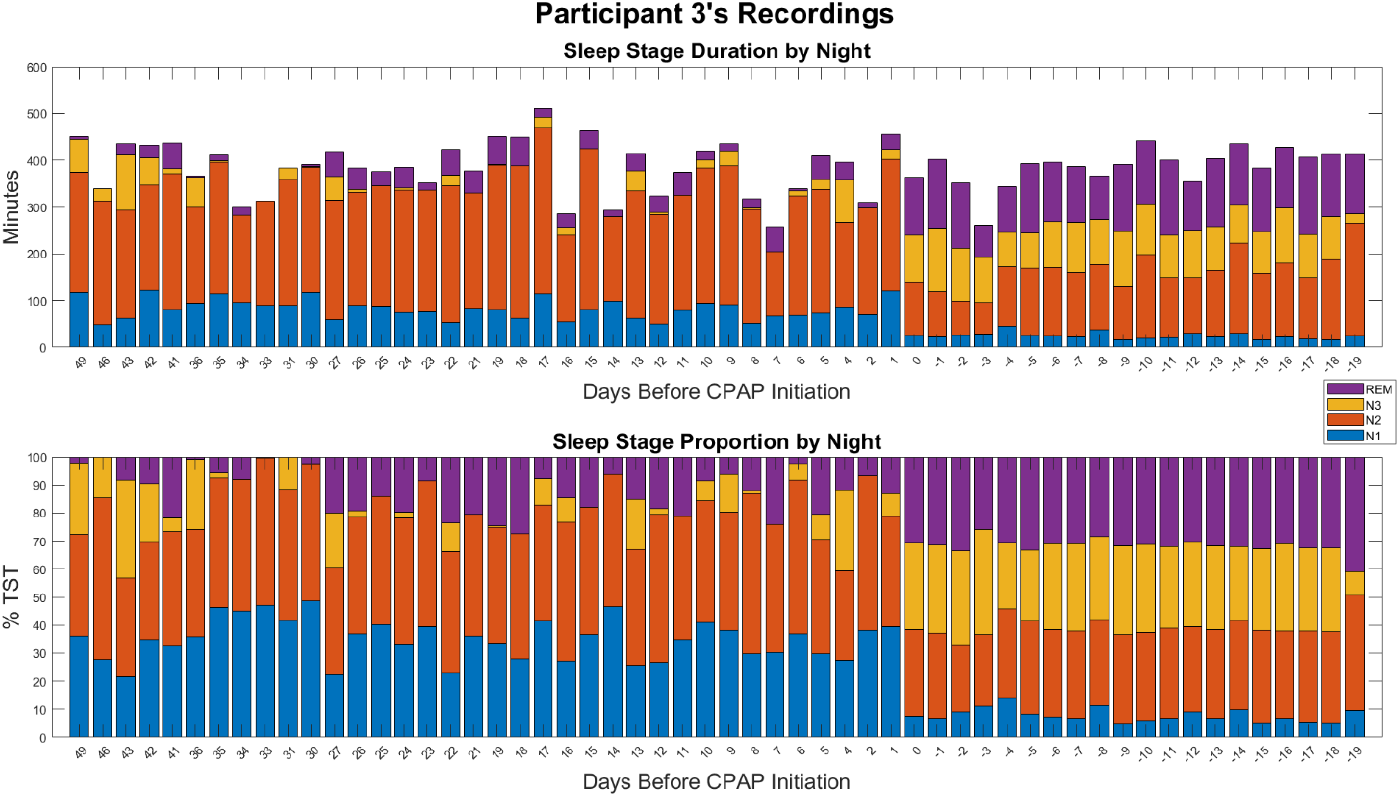
Bar chart displaying sleep stage duration and proportion for all usable recordings made by participant 3, in relation to CPAP initiation date.

## Discussion

We found strong differences of change in sleep architecture and features (micro arousals, awakenings and sleep stage shifts), Pre-CPAP and Post-CPAP using low-density EEG headbands for multi-night testing at home during routine diagnosis and treatment for OSA. The reduction in N1 and N2 and increase in N3 and REM, Post-CPAP is consistent with previous research using a full in laboratory polysomnography ^1–3,11^. Additionally, the decrease in micro arousals is also consistent with prior work ^11^, while the decrease in awakenings and stage shifts Post-CPAP have not previously been reported.

Strength testing showed that one participant had a strong influence on the overall group effects of our measures, and changes in N1,N2, N3 and awakenings were no longer significant once this participant was removed. This is likely due to participant 3 having the most severe OSA in our cohort. However, the direction of effects remained consistent for all measures except for N3 proportion. The increase in REM and decreases in micro arousals and sleep stage shifts Post-CPAP initiation retained significance when removing any single participant (including pt 3) and can be interpreted as robust findings.

To our knowledge there is only a single study using multi-night assessments in the short and medium term to test how sleep features in patients with severe OSA differ from controls at baseline and across multiple nights of CPAP use. This study reported that the duration of both N3 and REM increased sharply during the first night of CPAP, changing from less than to greater than control levels, but also that these measurements subsequently reduced linearly, to where they were no longer different to control levels by night 30 ^11^. These results confirm that at least in the case of severe OSA sleep recovery occurs over time and night-to-night variability is an important consideration when assessing the restorative properties of CPAP. Our results suggest that the changes in sleep architecture may not be so broad in people with less severe OSA.

OSA has a broad clinical manifestation ^10,20^, and it is extremely unlikely that OSA disrupts all sleep features in the same way across distinct patient groups. For instance Ye at al showed that within patients with at least moderate OSA, only 33% were clustered as being in a disturbed sleep group, while 25% were grouped as minimally symptomatic and 43% as excessively sleepy. However despite these phenotypic differences within the general OSA population, and our broad inclusion criteria for this study, we have been able to identify improvements in sleep quality over the entire group that were robust to strength testing. Although not all sleep features were significant without the inclusion of our very severe patient. A better understanding of specific impairments in sleep and their restoration in different subgroups of OSA (which may be more nuanced in non-severe cases), is a vital challenge for the progress of personalised OSA care. Because without clear understanding of the mechanistic disruption during sleep, which drives daytime impairment, it is not possible to reliably develop more specified treatment delivery programmes for distinct subgroups. Our data suggest that portable headbands are likely a useful tool to be able to investigate these challenges, firstly by avoiding the problem of night-to-night variation, and secondly facilitating large-scale data collection at a relatively low cost.

The unbalance data acquired across participants during this study is both a strength and limitation. It can be seen as a success that we have been able to collect 144 high-quality EEG recordings in the community, during a normal clinic timeframe without disrupting patient care. It is concerning that we lost 35% of recordings due to bad quality, however losing sessions due to bad signal is not uncommon in research using full polysomnography data, for instance the D’Rozario study excluded 40 participants due to excessive EEG artefact ^1^, and it is considerably more troublesome losing a polysomnography session due to poor quality than it is a portable device. As in-hospital studies require more resources, and recordings with portable devices can easily be re-attempted the next night, possibly after a virtual consultation with a technician if required.

The variability in the number of successful recordings between our most active and least active participant was sizeable (52). This can be addressed in research through statistical tools like mixed-effects modelling and strength testing, as demonstrated here. However if portable EEG are to become a tool to monitor patients for clinical motives, more research should be done to better understand the specific issues patients might encounter when conducting multi-night testing, and identify strategies to improve effective usage.

In our study, a larger number of trials were collected in the Pre-CPAP condition, although for participants who contributed data to both conditions, we retained higher percentage of Post-CPAP recordings (64.6% Pre-CPAP, 74.1 % Post-CPAP). This may reflect having a longer monitoring period in the Pre-CPAP condition (which varied between participants due to our design) and the more stable sleep patterns observed Post-CPAP, with less awakenings and shifts which could improve the likelihood of recordings meeting our required quality thresholds. However this may also reflect some degree of study fatigue, order effects using the EEG device or uncertainty when commencing a novel treatment programme. Notably two participants recorded more sessions after CPAP initiation, and it is extremely encouraging that the headbands can be used to record good quality data alongside CPAP.

## Conclusion

We have shown that portable low-density EEG headbands used at home are able to measure changes in sleep quality Pre-CPAP and Post-CPAP. This method may be a suitable for evaluating sleep quality across OSA subgroups over an extended periods, addressing night-to-night variation. Future work should test if specific changes or lack of changes in sleep quality during the early stages of CPAP initiation are predictive of longer-term restoration of daytime symptoms, and explore how CPAP might have a different effect on sleep for distinct severity groups and clinical phenotypes of OSA, as outlined in other work ^10^. Beyond this, we also see potential for this methodology to be combined with ever more accessible CPAP compliance data, to better understand the implications of different patterns of compliance (e.g. the influence of occasional missed nights), and how this impacts sleep quality and real-world performance, such as driving.

## Data Availability

At the time of submission all data produced in the manuscript is unavailable, as required by our ethical authorisations.

## Acknowledgements

We would like to express appreciation to all the participants who volunteered to take part in the study, Royal Papworth Charity for financial support, and all departments across the University of Cambridge and Royal Papworth Hospital for facilitating study set-up and delivery.

